# Pharmacokinetic analysis of intermittent rapamycin administration in early-stage Alzheimer’s Disease

**DOI:** 10.1101/2025.03.18.25324017

**Authors:** Helen Annervik Wallgren, Miia Kivipelto, Pontus Plavén-Sigray, Jonas E. Svensson

## Abstract

**Background:** rapamycin, an mTOR inhibitor used clinically for immunosuppression, shows promise for repurposing in age-related disorders including Alzheimer’s disease (AD). While the pharmacokinetics of daily rapamycin are well-characterized in transplant populations, limited data exist on intermittent dosing regimens in patients with neurodegenerative conditions.

**Method:** this open-label pilot study investigated the pharmacokinetic properties of weekly oral rapamycin in 13 patients with early-stage AD. Participants received 7 mg weekly (11 patients) or reduced doses (2 mg and 4 mg; 2 patients) for 26 weeks. Blood concentrations were measured at four timepoints (pre-dose/Cmin, and 1-, 3-, and 48-hours post-dose) during week 13.

**Results:** moderate interindividual variability was observed across timepoints (coefficient of variation was 0.28-0.40), with the 48-hour sample showing the lowest variability (CoV = 0.28) and strongest correlation with Cmin from the previous dosing (r = 0.72). Estimate of terminal half-life (68.9 ± 13.6 hours) aligned with previous studies.

**Conclusions:** blood concentrations at Cmin were below immunosuppressive levels in all participants. Our findings suggest that weekly rapamycin administration in AD patients results in acceptable pharmacokinetic variability, supporting fixed-dose regimens in future trials. The 48- hour post-dose measurement appears optimal for monitoring blood concentrations. Additionally, our investigation into blood-brain barrier permeability revealed methodological challenges in directly measuring rapamycin in cerebrospinal fluid due to analytical sensitivity limitations. The foremost limitation of this study was the sparse blood sampling schedule, with Cmin collected from the previous dosing occasion which prevented a complete AUC-calculation.

## Introduction

Rapamycin (also known as sirolimus), an mTOR inhibitor and immunosuppressive drug, has been in clinical use for nearly three decades in preventing rejection after organ transplantation (1). During the early 2000s, a series of experiments showed that rapamycin prolonged the lifespan in several model organisms, including yeast, C. elegans, and the fruit fly Drosophila melanogaster (2–4). In 2009 treatment with rapamycin was first shown to prolong lifespan of mice (5). These findings have since been replicated repeatedly (6,7) and subsequent animal research has shown a positive effect of rapamycin treatment on several age-related conditions, including neurodegenerative disease, such as in transgenic mouse models of Alzheimer’s disease (8,9).

These promising pre-clinical results have led to calls for repurposing rapamycin, and structurally similar mTOR inhibitors known as “rapalogs”, as treatments in a range of age-related diseases, with several studies already underway. Human clinical trials have been conducted to assess potential benefits of rapamycin and rapalogs in multiple age-related conditions, such as immunosenescence and decline in lean tissue mass (10–13) with more trials ongoing (14). Other trials have focused on neurodegenerative diseases, such as amyotrophic lateral sclerosis (15), multiple system atrophy (16), multiple sclerosis (17), and Alzheimer’s disease (clinicaltrial.gov ID: NCT04629495).

When rapamycin is used for immunosuppression, therapeutic drug monitoring (TDM) using whole blood trough values is recommended (18). The reason for this is a relatively narrow therapeutic window, together with an substantial observed interindividual variation in bioavailability and elimination (19). It is unclear whether the interindividual variation presents the same challenge in new non-immunosuppressive indications where other dosing regimens have been used (11,13,17). Therefore, the question of whether TDM is necessary or if fixed dosing would be sufficient remains to be determined.

For treatment in new indications where immunosuppression is not desired, it has been suggested to administer rapamycin at lower doses but also using an intermittent dosing regimen. These proposals are based on rapamycin’s specific binding profile to subtypes of the mTOR complex, where it has been hypothesized that intermittent dosing could reduce side effects while maintaining treatment efficacy (14). Whereas the pharmacokinetics of daily dosing of rapamycin is well described in healthy populations and in patients following organ transplantation, less is known about the pharmacokinetic properties in other groups such as patients with neurodegenerative disease. Additionally, human data on rapamycin’s pharmacokinetics under intermittent dosing remain scarce, as does evidence regarding its ability to cross the blood-brain barrier (BBB).

Here we investigate the pharmacokinetic properties of oral tablet rapamycin administered once weekly to patients with early Alzheimer’s disease. We aim to:

1. Provide data on the interindividual variability in blood concentration in order to guide decisions on whether to use a fixed dose or TDM in future rapamycin-trials applying intermittent dosing regimens.
2. Assess the optimal time point for blood concentration monitoring during intermittent dosing.
3. Provide data on rapamycin’s pharmacokinetic properties during long-term intermittent treatment, including BBB permeability.

## Methods

### Clinical trial design

The study is part of a single-center, open-label, one-arm pilot clinical trial where treatment with rapamycin was evaluated in patients with early-stage Alzheimer’s disease (20). The study was conducted in accordance with the Declaration of Helsinki and the International Conference on Harmonisation for Good Clinical Practice (ICH GCP) and was approved by the Swedish Medical Products Agency (5.1–2023-8283), and the Swedish Ethical Review Authority (2023–03075-02). Before study start, the trial was registered at ClinicalTrials.gov (NCT06022068) and EudraCT (2023–000127-36). Written informed consent was collected from all participants and their designated study partners before initiating any study procedures.

The study was conducted as a collaboration between Karolinska Institutet and the Karolinska University Hospital, Theme Inflammation and Aging. Patients followed at the Solna Memory clinic were eligible for inclusion. A full description of inclusion and exclusion criteria has been published elsewhere (20). In short, patients between 50-80 years, diagnosed with amyloid-positive Alzheimer’s disease, either with mild cognitive impairment (MCI) or early-stage Alzheimer’s dementia (not above stage 4 according to the National Institute of Aging-Alzheimer’s Association 2018 criteria) (21), and with a normal or clinically acceptable medical history and physical examination were eligible to participate in the trial. All study participants underwent routine laboratory testing and brain imaging with MRI prior to the first dose of the study drug. Throughout the study, participants were continuously monitored for safety and adverse events through regular clinical follow-up visits, including safety blood tests.

### Dosing regimen and sampling procedures

Participants were administered a weekly dose of rapamycin for 26 weeks, starting with an initial dose of 3 mg, which, if well tolerated, was increased to the target dose of 7 mg in the second week. Participants were prescribed 1 mg tablets of rapamycin (Tablet Rapamune®) which they collected at the hospital pharmacy. A study nurse dispensed the drug into a pill organizer which participants were asked to bring to clinical check-ups where it was verified that the tablets were taken according to the study protocol.

At the mid-trial follow-up visit (week 13), blood samples for sirolimus concentration were collected at four time points over 48 hours. Blood concentration tests were taken just prior to the weekly dose (C_min_), and at 1-, 3- and 48-hours post-dose. The 1h sample was chosen with the aim of capturing C_max_ based on estimates from prior studies (22,23). Participants were fasting at the time for the pre-dose test due to simultaneous sampling of routine laboratory variables. Following the blood draw the participant was instructed to swallow the weekly dose with a glass of water and received immediately thereafter a light meal. The pre-dose, 1- and 3-hour blood samples were collected by a study nurse as part of the study visit. The participants were given the option to select a caregiver closer to their home for the 48-hour blood test. For assessment of BBB permeability of the study drug, a lumbar puncture for collection of a cerebrospinal fluid (CSF) sample was performed before initiation of the study treatment and again within 28 days of the last dose.

Sirolimus whole blood concentrations were analyzed at Karolinska University Hospital Laboratory, Huddinge, Medical Unit Clinical Pharmacology, a unit which routinely performs clinical sirolimus concentration analyses. Analysis was performed on a Thermo Fisher (Thermo Scientific, Waltham, MA, USA) liquid chromatography–tandem mass spectrometry (LC-MS/MS) machine. All blood samples were collected, stored and transported as per clinical routine outlined in written instruction supplied by the laboratory.

### Blood brain-barrier-permeability analysis

At steady state the CSF concentration of a drug that passes freely over the BBB can be expected to be similar to the free concentration in blood (24). Given that the free fraction of rapamycin is only about 0.17% of whole blood (25), it is uncertain whether standard laboratory methods can reliably quantify it in CSF. The lower limit of quantification (LLOQ) for rapamycin in whole blood using LC-MS/MS—the gold standard method—typically ranges from 0.5 to 1 ng/mL (26), which is more than an order of magnitude higher than expected CSF concentrations at therapeutic doses. To determine whether lower CSF concentrations of rapamycin can be quantified, we conducted a method assessment as part of this pharmacokinetic study. Rapamycin (purity > 98%, Thermo Scientific Chemicals) was used to prepare calibration standards in blank CSF (i.e., CSF from individuals not treated with rapamycin). The study was performed at Drug Discovery and Development Platform, SciLifeLab in Uppsala, using a triple quadrupole mass spectrometer (Xevo™ TQ-S micro, Waters Corp.), with the aim of determining the LLOQ of rapamycin in CSF. Further methodological details are provided in the Supplementary Materials.

### Pharmacokinetic modeling and statistical analysis

The interindividual variation in blood concentration of rapamycin was assessed through calculating the standard deviation (SD) and the coefficient of variation (CoV) at each of the sampling times separately (C_min_, 1h, 3h, and 48h tests). Since the C_min_ sample was collected from a separate dosing occasion a complete area under the curve (AUC) of the blood concentration was not possible to calculate. The concentration data collected post dosing was performed at 1h, 3h and 48h, allowing for calculation of AUC_0-48h_. However, given the relatively sparse sampling scheme with only one sample after the distribution phase, together with the substantially higher concentrations in the two early samples compared to the 48h sample, the observed variation in AUC_0-48h_ values will be mainly influenced by the variation in the 3h sample. Consequently, this makes it not a valid measure of the true intraindividual variation in AUC.

In order to estimate terminal half-life (terminal t½), more than one concentration sample collected after the distribution phase (i.e., during the elimination phase) of the drug is needed. To overcome this, we made the assumption that C_min_ is stable between doses and hence used C_min_ from the previous dosing occasion to provide an additional time point (denoted 168h below). To underscore that the terminal half-life calculation was based on blood concentration values from two separate dosing occasions this parameter is denoted as “*pseudo-*” terminal t½ below, and it was calculated as:

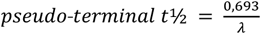

where λ denotes the terminal elimination rate constant, calculated as the negative slope of the logarithmed concentration versus time for the two last time points (i.e., 48h and 168h).

In order to assess the association between the different post-dosing samples, and the stability over time, Pearson’s correlation coefficient was calculated between C_min_ from the previous dosing occasion and concentration estimates at the sampling times post-dose i.e., at 1h, 3h, and 48h. The following heuristics were used to describe the strength of association between variables: a correlation estimate between 0–0.19 was described as negligible, 0.2–0.39 as weak, 0.40–0.59 as moderate, 0.6–0.79 as strong and 0.8–1 as very strong (25).

Pharmacokinetic calculations were performed using Microsoft Excel (Microsoft Corporation, Redmond, WA, USA). Data visualization and additional statistical analyses were conducted using R software (version 4.2.2, R Foundation for Statistical Computing, Vienna, Austria).

## Results

14 patients, eight women and six men with an average age of 60.9 ± 4.2 (mean ± SD) years were included in the study. Six patients had a diagnosis of MCI and eight of dementia, with mean time since diagnosis of 6.6 ± 4.2 months and Montreal Cognitive Assessment (MoCa) score of 23.8 ± 3 (maximum 30) at inclusion. The participants’ mean weight was 75.5 ± 16 kg with body surface area of 1.9 ± 0.2 m^2^.

One participant (male) did not tolerate the study drug due to nausea and was excluded from the trial two weeks after initiating the treatment (i.e., before sampling of blood concentration). Consequently, the results are based on the remaining 13 participants, who all completed the 26-week treatment. Two participants (both female) received a lower dose of rapamycin (2 and 4 mg respectively). The reason for the reduced dose was side effects in one participant (vertigo following initial dose) and safety reasons (chronically elevated aspartate transaminase) for the second participant. For both participants the treatment was well tolerated at these doses. Three participants did not perform the 3h sample, one of whom also provided blood sample at 24h instead of 48h. All participants were on a stable dose of a cholinesterase inhibitor (minimum of 4 weeks of target dose before initiation of study drug). No serious adverse events were observed during the trial.

Average concentration values from participants taking 7 mg rapamycin (11 out of 13) at the different timepoints are reported in Table 1, together with the results from the two participants taking reduced doses. For participants receiving full dose C_min_ was 1.17 ± 0.44 ng/mL (CoV = 0.38), and 1h after dosing 26.73 ± 10.74 ng/mL (CoV = 0.4).

**Table 1.**
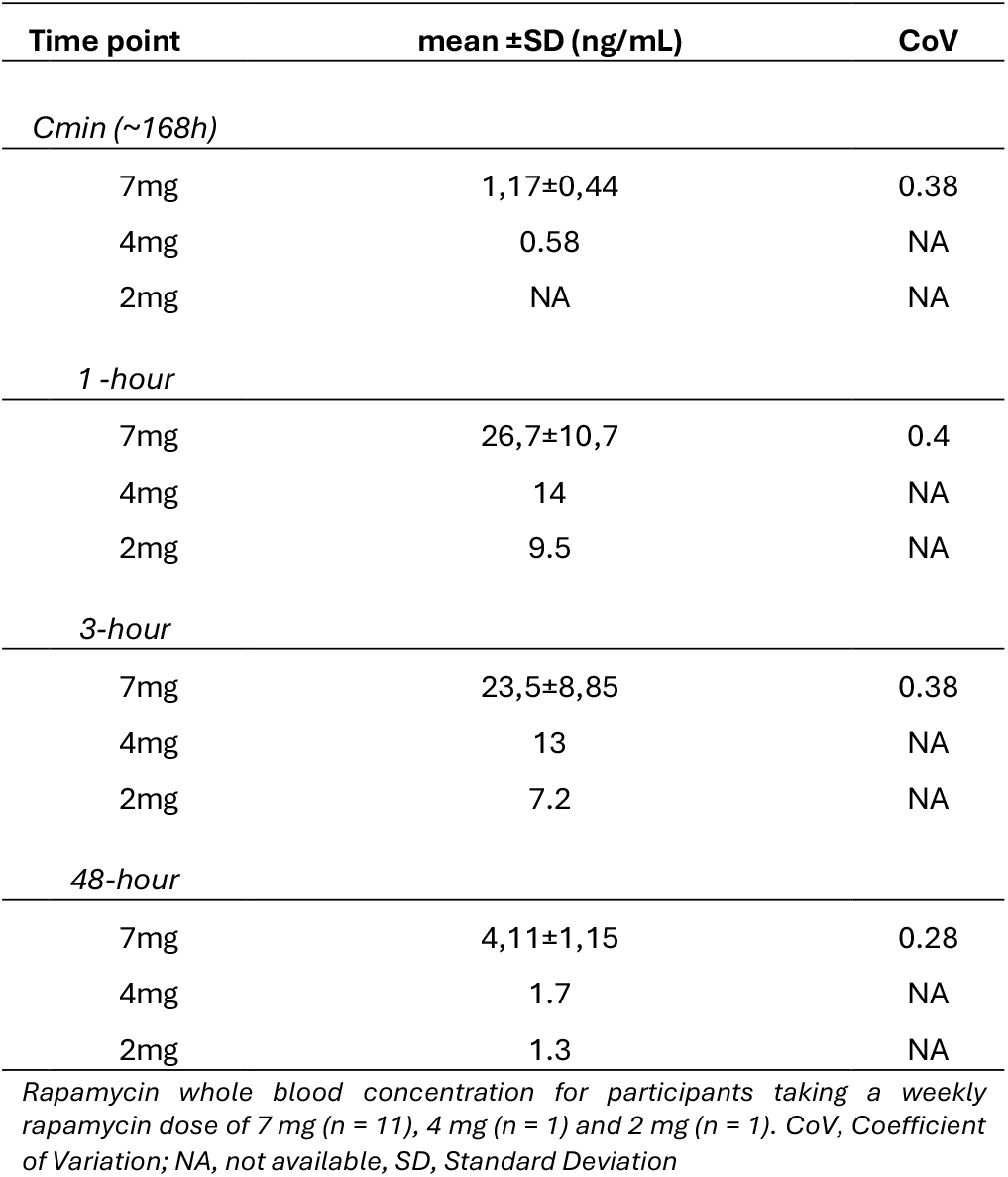
Rapamycin whole blood concentration.

| Time point | mean ±SD (ng/mL) | CoV |
| --- | --- | --- |
| *Cmin (~168h)* |  |  |
| 7mg | 1,17±0,44 | 0.38 |
| 4mg | 0.58 | NA |
| 2mg | NA | NA |
| *1 -hour* |  |  |
| 7mg | 26,7±10,7 | 0.4 |
| 4mg | 14 | NA |
| 2mg | 9.5 | NA |
| *3-hour* |  |  |
| 7mg | 23,5±8,85 | 0.38 |
| 4mg | 13 | NA |
| 2mg | 7.2 | NA |
| *48-hour* |  |  |
| 7mg | 4,11±1,15 | 0.28 |
| 4mg | 1.7 | NA |
| 2mg | 1.3 | NA |

*Rapamycin whole blood concentration for participants taking a weekly rapamycin dose of 7 mg (n = 11), 4 mg (n = 1) and 2 mg (n = 1). CoV, Coefficient of Variation; NA, not available, SD, Standard Deviation*

Correlation analysis of blood concentrations at different time points for participants taking 7 mg per week are reported in Figure 2. The C_min_ value had the strongest correlation to the 48h sample (r = 0.72) and the weakest correlation to the 1h sample (r = 0.076). A very strong correlation was seen between the 1h and 3 h sample (r = 0.87) and between the 3h and 48h sample (r = 0.80).

**Figure 1.**
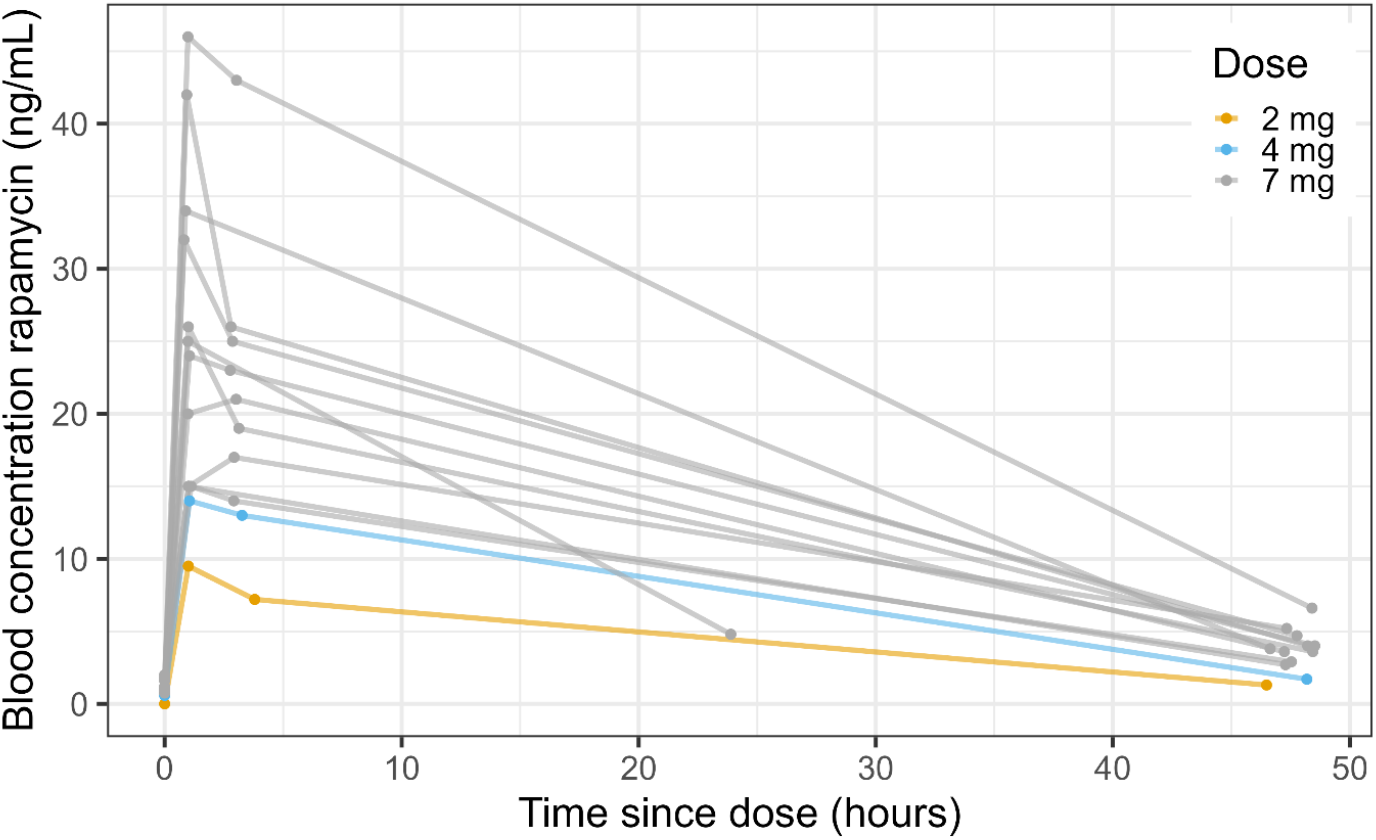
Rapamycin whole blood concentration before intake of weekly dose (C_min_) and at three different time points following dosing (1, 3 and 48 hours post dose, with one participant tested at 24h instead of 48h post dose). Data from participants on a weekly dose of 7 mg in gray, with the participants on 2 and 4 mg doses shown in yellow and blue respectively.

**Figure 2.**
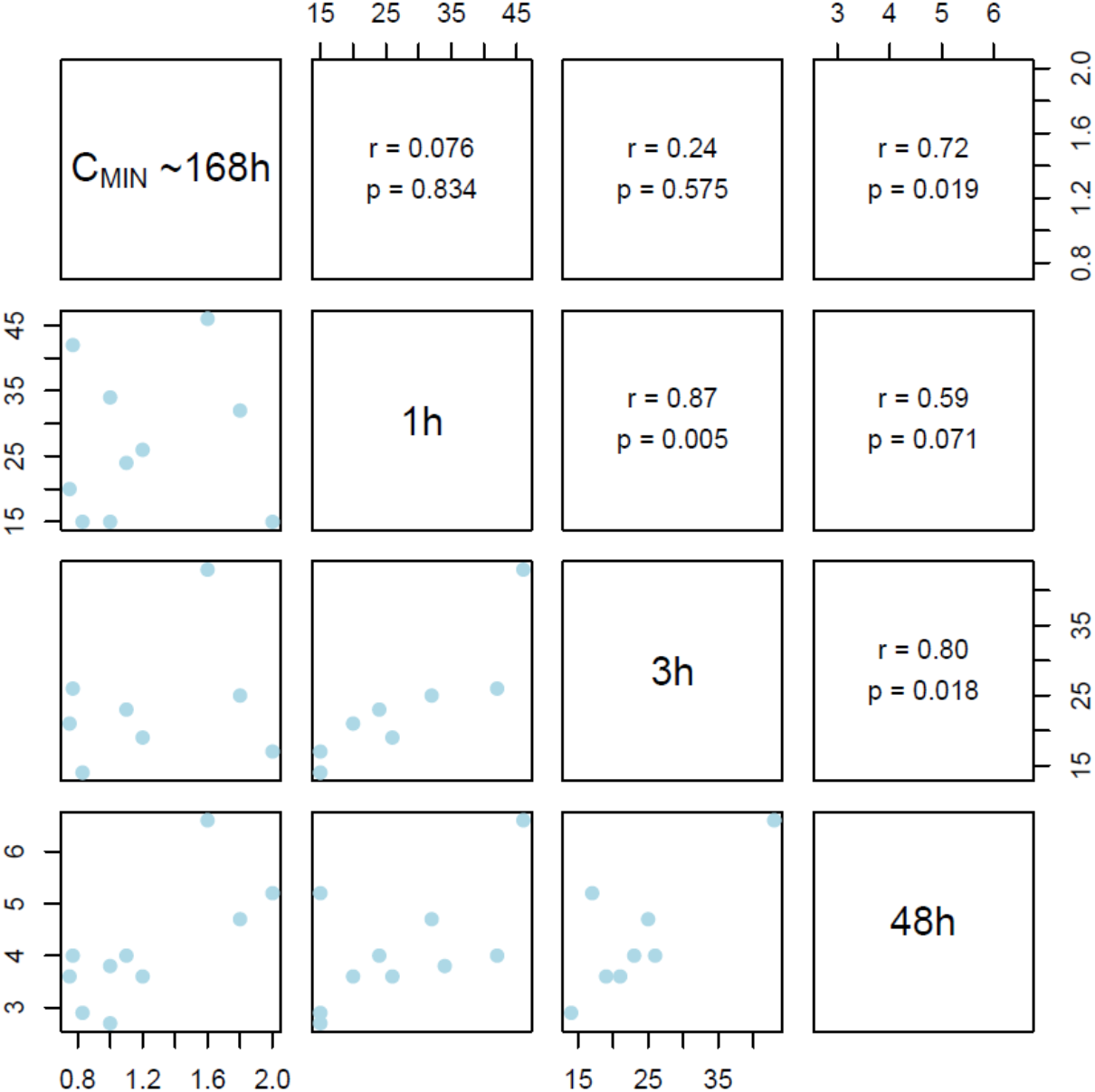
Correlation estimates for rapamycin whole blood concentration before intake of weekly dose (C_min_) and at three different time points following dosing (1, 3 and 48 hours post dose). Only participants taking 7mg per week is included in this analysis.

For participants receiving 7 mg, pseudo-terminal t½ was 68.85 ± 13.64 hours (CoV 0.20). The participant missing the 48-hour sample was not included in this calculation. The participant receiving a reduced dose of 4 mg showed a pseudo-terminal t½ of 77.3 hours and for the participant receiving 2 mg we could not estimate pseudo-terminal t½ due to a non-quantifiable blood concentration at C_min_.

For method development of rapamycin quantification in CSF, an LLOQ of 20 ng/mL was achieved. Sample pre-concentration did not significantly enhance sensitivity, yielding an LLOQ of 10–20 ng/mL. See Supplementary Material for further details.

## Discussion

The purpose of the study was to characterize the pharmacokinetics of rapamycin when given weekly to patients with early Alzheimer’s disease. The drug is of high interest in the field of aging research, with multiple ongoing studies evaluating its safety and efficacy for new indications (14). However, limited data on the pharmacokinetic properties for long-term, intermittently administered rapamycin exist, with only a few studies evaluating the topic (26,27). In our study, blood concentrations showed moderate interindividual variance, with CoV ranging from 0.28 to 0.40 across timepoints. The 48h sample demonstrated the lowest variance (CoV = 0.28) and the strongest correlation with C_min_ from the previous dosing (r = 0.72), suggesting 48h as the time point of choice for control of blood concentration based on this series of samples.

When rapamycin is used for immunosuppression following organ transplantation, TDM using C_min_/trough values is recommended based on two key factors: the drug’s narrow therapeutic window in this indication (5-15 ng/mL) and a substantial observed interindividual pharmacokinetic variability (19). At blood concentrations below 5 ng/mL, immunosuppression may be insufficient, while levels exceeding 15 ng/mL increase the risk of adverse effects and toxicity. For age-related disorders where immunosuppression is not desired, intermittent and/or lower dosing of rapamycin and rapalogs is typically employed (10–13), requiring reassessment of these factors. In our study, the 11 patients receiving weekly 7 mg doses, blood concentrations at 48h averaged 4.11 ± 1.15 ng/mL (range: 2.7–6.6 ng/mL, CoV = 0.28), with only two participants exceeding 5 ng/mL. Notably, no participants exceeded 5 ng/mL at C_min_. While earlier time points showed moderately higher variability (CoV = 0.38–0.40), this still remains lower than that observed in transplant patients on rapamycin treatment (28) as well as with specific commonly used drugs such as certain beta blockers (29) and antidepressants (30).

Although the minimum effective concentration for indications other than immunosuppression remains unknown, our findings demonstrate that weekly dosing avoids concentration levels that could be of high safety-concern, while exhibiting relatively low pharmacokinetic variability, supporting a fixed-dose approach in future trials.

No recommendations exist for the optimal timing of testing the blood concentration of rapamycin when administered as a weekly dose. Potential sampling points include C_max_, C_min_, or at some earlier time during the elimination phase (i.e., between the end of the distribution phase and C_min_). In daily dosing of rapamycin, C_min_ has been shown to have an excellent correlation to AUC (23,28). In our data, based on a weekly dosing scheme of 7 mg rapamycin, values of C_min_ are low (0.75–2 ng/mL), approaching the LLOQ of 0.5 ng/mL at our hospital lab. Given a lower dose or the use of an analysis method with higher LLOQ, it is likely that some samples would not be able to be quantified, as demonstrated by our 2 mg dose participant’s unmeasurable C_min_. Time to peak concentration (T_max)_ for orally administered rapamycin is reported at approximately 1h, although some studies indicate considerable interindividual variation, with T_max_ ranging from 0,5 to 3h (19). In our data, 11 participants showed the highest concentration at 1h, however, two participants peaked at the 3h sample. This variation complicates reliable C_max_ capture with single-point sampling, reflected by the fact that the highest variation (CoV = 0.40) in our data was at the 1h timepoint. Both C_min_ and 3h samples showed CoV of 0.38, while the 48h sample demonstrated lowest variation (CoV = 0.28). Taken together, our results suggest that sampling early during the elimination phase is the best option when rapamycin is administered as a weekly dose.

In our data, the long terminal half-life (pseudo-terminal t½) of 68.85 ± 13.64 hours is well in line with that observed in multiple previous studies evaluating pharmacokinetic properties after a single dose rapamycin in healthy volunteers, as well as after multiple doses in transplant recipients (22,23,31). The two participants treated with reduced doses (2 and 4 mg) showed consistently lower blood concentrations across all time points, with the 2 mg dose producing the lowest results, suggesting dose proportionality for rapamycin administered once per week within this dose range.

The extent to which rapamycin passes the BBB is unclear. The drugs high molecular weight (914.2 Da) and its status as an efflux pump substrate suggest limited permeability. One study of rapamycin in patients with amyotrophic lateral sclerosis showed no quantifiable levels of sirolimus in CSF using LC-MS/MS (15). To assess whether LC-MS/MS could reliably quantify rapamycin in CSF, we developed a method achieving an LLOQ of 20 ng/mL. While further optimization might lower this threshold, reaching the single-digit pg/mL range - necessary to definitively rule out the presence of a pharmacologically relevant concentration - was deemed unfeasible. Given this, we elected not to perform any analysis of rapamycin concentration on the CSF data collected from the study participants in our trial. This constrained LLOQ should also be taken into consideration when interpreting the published dataset where no rapamycin could be detected in CSF of patients on the drug (15). While some clinical data suggest BBB passage— such as rapamycin’s efficacy in treating cerebral manifestations of tuberous sclerosis complex in humans (33)—the question of its extent of brain delivery remains open. Given the expected low free concentration in CSF, one option for future research is to perform post-mortem analysis of brain tissue from rapamycin-fed laboratory animals. Alternatively, labelling rapamycin with a radioactive nuclide and perform micro-dosing experiments using positron emission tomography (PET) could provide further insights into its brain exposure.

The foremost limitation of this study was the sparse blood sampling schedule, with C_min_ collected from the previous dosing occasion. This prevented a complete AUC estimation and limited the analysis in two ways: 1) we could not evaluate interindividual variation in overall drug exposure but had to restrain the analysis to the separate sampling time-points; 2) more than one concentration sample collected after the distribution phase of the drug is needed to perform a full modelling of the drug’s pharmacokinetic parameters, such as t1/2 . To overcome this, we assumed stable C_min_ between doses and used the previous dosing occasion’s C_min_ as an additional timepoint, which allowed for estimates of pseudo-terminal t½.

In conclusion, our data show that weekly rapamycin administration in patients with early Alzheimer’s disease results in moderate interindividual variability in blood concentration, suggesting that a fixed dosing regimen may be appropriate for future trials using a similar design. Of the four time points assessed (C_min_, 1h-, 3h-, and 48h-hours post dose) the 48h sample showed the lowest interindividual variability and strongest correlation to C_min_ from the previous dose, supporting it as the optimal sampling time for monitoring blood concentration.

## Data Availability

All data produced in the present study are available upon reasonable request to the authors and pending approval from the Karolinska Institutet Data Compliance Office.

## List of abbreviations

AUC: Area under the curve
BBB: Blood-brain barrier
CoV: Coefficient of variation
CSF: Cerebrospinal fluid
LC-MS/MS: Liquid chromatography–tandem mass spectrometry
LLOQ: Lower limit of quantification
MCI: Mild cognitive impairment
SD: Standard deviation
TDM: Therapeutic drug monitoring

## Declarations

### Ethics approval and consent to participation

The study was approved by the Swedish Medical Products Agency (5.1–2023-8283), and the Swedish Ethical Review Authority (2023–03075-02). Written informed consent was collected from all participants and their designated study partners before initiating any study procedures.

### Availability of data and materials

The datasets used and analyzed during the current study are available from the corresponding author on reasonable request.

### Authors’ contributions

JS, PPS and MK conceived of and designed the study. PPS and MK directed and supervised conduction of the study. JS and HA performed the data collection and drafted the manuscript. JS, HA and PPS performed data analysis. All authors contributed to the interpretation of the results, critically reviewed the manuscript, and approved the final manuscript.

## Acknowledgements

We wish to thank Associate Professor Staffan Rosenborg, MD, PhD at the Clinical Pharmacology Unit, Karolinska University Hospital Huddinge for valuable discussions on the blood sirolimus analysis method, and Associate Professor Anna Matton, PhD at the Karolinska Institute, for providing blank CSF. The authors would like to acknowledge support of the SciLifeLab Drug Discovery and Development Platform, Sweden.

