## Supplementary Information for "Pharmacokinetic analysis of intermittent rapamycin administration in early-stage Alzheimer’s Disease"

### Preparation of Standards and Sample

A nine-point calibration curve was prepared in blank human cerebrospinal fluid (CSF), with rapamycin concentrations ranging from 0.5 nM to 50 nM (approximately 0.5–50 ng/mL). Aliquots of 20 µL from each calibration standard were diluted with 80 µL of ice-cold methanol containing 50 nM warfarin as an internal standard. The samples were processed in a 96-well plate and centrifuged at 2465 × *g* at 4 °C for 20 minutes. A 5 µL aliquot from each well was injected into the LC-MS/MS system for analysis.

Following initial analysis, samples were evaporated to dryness and reconstituted in 20 µL methanol/water (1:1, v/v). A 10 µL aliquot was subsequently injected into the LC-MS/MS system.

### Chromatographic Method

Chromatographic separation was performed on a BEH $C_8$ column (2.1 × 50 mm, 1.7 µm) at a controlled temperature of 60 °C. The mobile phases consisted of solvent A (10 mM ammonium formate in water) and solvent B (10 mM ammonium formate in methanol), both adjusted to pH 3.3.

Gradient elution was applied as follows: an initial composition of 10% B was maintained from 0.00 to 0.50 min, followed by a linear increase to 100% B at 1.20 min. This composition was held constant until 1.90 min, after which the gradient returned to 10% B at 2.00 min. The flow rate was set at 0.5 mL/min. A 10 mM ammonium buffer was necessary to effectively displace $Na^+$ and $K^+$ ion adducts, allowing exclusive formation of ammonium adducts, which are susceptible to fragmentation in the collision cell, enabling multiple reaction monitoring (MRM) detection.

### Mass Spectrometry Conditions

Mass spectrometric detection was performed using a Xevo™ TQ-S micro triple quadrupole mass spectrometer (Waters Corp.) operating in positive electrospray ionization ($ESI^+$) mode. The instrument was set to MRM mode, with the following optimized parameters: capillary voltage of 3 kV, desolvation gas temperature of 100 °C, and a desolvation gas flow rate of 1000 L/h.

For quantitative analysis, MRM transitions were optimized for warfarin and rapamycin, including parent and fragment ions, cone voltages (Cone), and collision energies (CE). The warfarin transition was monitored at m/z 309.1596 → 163.0050, with a cone voltage of 28 V and a collision energy of 16 V. The MRM transitions for rapamycin were systematically optimized as follows: 931.37 → 351.13 (Cone: 5 V, CE: 25 V, quantifier ion), 931.37 → 452.09 (Cone: 10 V, CE: 30 V), 931.37 → 474.06 (Cone: 5 V, CE: 25 V), 931.37 → 492.08 (Cone: 10 V, CE: 20 V), 931.37 → 510.03 (Cone: 10 V, CE: 25 V), 931.37 → 524.06 (Cone: 10 V, CE: 30 V), 931.37 → 542.07 (Cone: 5 V, CE: 25 V).

### Conclusions

Rapamycin is a cyclic compound with a molecular weight of 914.2 Da. During electrospray ionization (ESI), it predominantly forms $Na^+$ and $K^+$ adducts, which are resistant to collision-induced fragmentation (CID), making MRM-based detection challenging. This limitation was addressed by introducing ammonium-based buffers into the mobile phases to effectively displace metal ions. Additionally, the desolvation temperature was maintained at 100 °C to retain ammonium ions in solution. As a result, a stable rapamycin-ammonium ion complex (m/z 931.37) was generated, facilitating the development of a robust MRM method.

Using a standardized protein precipitation method with an organic solvent and a 2-minute LC-MS/MS method, an LLOQ of 20 nM (≈ 20 ng/mL) was achieved.

Given that the general sensitivity of the TQ-S micro system ranges from 0.5–10 nM, depending on the analyte and biological matrix, achieving lower LLOQ values may prove challenging with the current instrumentation.
